# Cerebrospinal fluid profile, including α-Synuclein seeding activity, of p.A53T SNCA mutation carriers: Data from the PPMI Study

**DOI:** 10.64898/2026.07.23.26356817

**Authors:** Athina Maria Simitsi, Nikolaos Papagiannakis, Ioanna Alefanti, Marina Picillo, Paolo Barone, David-Erick Lafontant, Kenneth Marek, Andrew Siderowf, Tanya Simuni, Christos Koros, Leonidas Stefanis, the Parkinson’s Precision Medicine Initiative

**Affiliations:** National and Kapodistrian University of Athens, 1st Department of Neurology, Eginition Hospital, Athens, Greece; Centre for Neurodegenerative Diseases (CEMAND), University of Salerno, Salerno, Italy, Department of Medicine, Surgery and Dentistry “Scuola Medica Salernitana”, University of Salerno, Salerno, Italy., Salerno, Italy; The University of Iowa, Iowa City, IA, United States of America; Institute for Neurodegenerative Disorders, New Haven, CT, United States of America; University of Pennsylvania, Neurology, Philadelphia, AL, United States of America; Northwestern University, Parkinson’s Disease and Movement Disorders Center, Northwestern University Feinberg School of Medicine, Chicago, USA, Chicago, IL, United States of America; IRCCS Synlab SDN, Naples, Italy

## Abstract

We report here, based on Parkinson’s Progression Markers Initiative (PPMI) data, on the Cerebrospinal Fluid (CSF) profile of a group of 12 Parkinson’s Disease (PD) subjects with the prototypical p.A53T SNCA mutation, comparing them to 36 matched Healthy Controls (HCs) and 36 idiopathic PD (iPD) cases. We furthermore assessed the CSF profile of 7 asymptomatic carriers of this mutation. There was no significant difference between the 3 groups of A53T-PD, HC and iPD in total alpha synuclein (a-syn) levels, β-amyloid 1-42, total-Tau and p-Tau, although A53T-PD subjects tended to have slightly lower β-amyloid 1-42, total-Tau and especially total a-syn levels. All A53T-PD cases had a positive CSF a-syn Seeding Amplification Assay (SAA). Four out of 7 asymptomatic carriers also had a positive SAA, in 3 without motor symptoms or signs and absence of clear prodromal manifestations. Conversion to motoric manifestations has occurred in one out of these 3 subjects, 8 years after SAA positivity, while one other subject only has hyposmia 8 years later. Overall, these results indicate that at least in early stages of PD, CSF Alzheimer’s Disease profiles are not significantly different in A53T-PD compared to HCs or iPD, while the CSF a-syn SAA is universally positive in this group. Furthermore, the assay may be positive in asymptomatic carriers at a time with no prodromal manifestations and many years before motor disease onset, opening a window into very early stages of disease pathobiology and opportunities for early therapeutic intervention.

## 1. Introduction

The p.A53T mutation in the alpha-synuclein (SNCA) gene represents an archetypal form of genetic synucleinopathy with many features of idiopathic Parkinson’s disease being exacerbated such as hyposmia, cognitive decline and RBD^1, 2, 3^.Parkinson’s disease (PD) as a concept is based on clinicopathological criteria, including Lewy-body (LB) pathology related to alpha-synuclein (α-Syn) aggregation^4^. However, some genetic forms of PD do not follow this rule, like *parkin* mutations that show nigral degeneration without LBs^5^ or LRRK2 mutations where the histopathology is variable ^6, 7^. Neuropathological studies in A53T-PD show severe and more widespread LB pathology than found in typical i PD^8^.

The newly reported method of detection of pathological α-synuclein (misfolded and aggregated) in CSF using the seeding amplification assay (SAA) has revolutionized research criteria in Parkinson’s disease^9^. Assessment of genetic forms of PD for a-syn seeding in the CSF is increasingly recognized as an important way to evaluate the underlying pathology.

The cerebrospinal fluid profile in terms of other conventional markers of neurodegeneration in the p.A53T SNCA cohort is still elusive. We have previously reported Alzheimer’s Disease-related biomarkers in a small cohort of PD patients harboring the p.A53T SNCA mutation^10^. We intend here to use Parkinson’s Progression Marker Initiative data to evaluate the cerebrospinal fluid profile of a p. A53T SNCA subgroup of study participants in terms of markers of neurodegeneration (including a-syn seeding with the SAA, alpha synuclein levels, Abeta amyloid 1-42, total-Tau and p-Tau), in order to achieve a more profound understanding of the pathophysiological features of this rare genetic synucleinopathy. In view of data in experimental transgenic mice overexpressing the pA53T human protein, that show very limited seeding activity using the wild type (WT) protein as substrate^11^, it is of special interest to ascertain whether the SAA, using the WT α-synuclein as substrate, is positive in such subjects with the heterozygote mutation, reflecting the severe LB pathology reported. If so and given the availability of longitudinal sample collection also from asymptomatic mutation carriers within PPMI, it would be important to ascertain the timing of SAA conversion from negative to positive relative to prodromal signs or symptoms, as well as motor disease onset; this could help identify very early stages of disease pathogenesis. Furthermore, given the abundant interest in copathologies, it will be important to establish whether there is any indication of the emergence of the classic neurodegeneration markers related to Alzheimer’s pathology in this rare genetic cohort.

## 2. Methods

### Data acquisition - Sharing

This is a retrospective observational cohort study and the data used in the preparation of this article were obtained on May 11^th^, 2026 from the Parkinson’s Progression Markers Initiative (PPMI) database (https://www.ppmi-info.org/access-data-specimens/download-data), RRID:SCR_006431. For up-to-date information on the study, visit http://www.ppmi-info.org. This analysis used mostly data openly available from PPMI, while data on asymptomatic carriers were obtained following special request. The present study was conducted in agreement with the principles of the Declaration of Helsinki and was approved by the Scientific Board of all PPMI sites involved. Signed informed consent was obtained from all participants recruited.

We collected from the PPMI database all A53T-PD with CSF data regarding a-syn seeding amplification assay (ASSAA), total a-syn (t-a-syn), Abeta amyloid 1-42, total-Tau and p-Tau (from now on called CSF-p) at Baseline visit (BL), all i-PD with data for CSF-p in all visits and all HC with data for total a-syn (t-a-syn), Abeta 1-42, t-Tau and p-Tau in all visits. We used propensity-based matching to identify i-PD matched with the A53T-PD for age at visit and age at diagnosis and HC matched with the A53T-PD for age at visit in a 3:1 ratio. Sex matching was not performed because the resulted original matching was statistically balanced between samples’ sex and avoided introducing further constraints. We also collected ASSAA CSF data for asymptomatic A53T mutation carriers (A53T-AC), both at BL and longitudinally, to explore the trajectory of the SAA over time, correlating with disease evolution.

We have collected from the PPMI dataset, except for the CSF data mentioned above, basic demographic characteristics (age at visit, age at diagnosis, and education) and basic clinical scores: MDS-UPDRS III-OFF state, Hoehn and Yahr scale (HCY) – OFF state, Montreal Cognitive Assessment (MOCA), Geriatric Depression Scale (GDS), UPSIT, RBDQS, SCOPA-AUT.

### Matching Procedure

Each participant – visit pair was considered as one observation for the purposes of matching. For A53T-PD participants, only their baseline visit was included, which was the basis for the selection, but there was no such constraint in the other categories.

A propensity score matching based technique was utilized to select the matching HC and iPD participants in a 3:1 ratio with the A53T-PD participants. The propensity score was computed through a logistic regression model with age and age of diagnosis (in the case of iPD only) as independent variables, and the presence of the p.A53T mutation as the dependent variable. Afterwards, a nearest neighbor matching algorithm was used to determine the matching participants. In case a participant was selected more than once with different visits, the earliest visit was kept, while their other visits were removed from consideration. The algorithm was rerun on the new participant–visit set, repeating the aforementioned process until no duplicates remained.

### Statistical analysis

Means and standard deviations were used to describe the values of all markers examined. For the baseline comparisons for normally distributed scores, One-way ANOVA was used for normally distributed scores while nonparametric Independent-Samples Kruskal-Wallis Test was used for non-normally distributed scores. For dichotomous data, analysis was performed using Fisher–Freeman–Halton Exact test. Pearson’s or Spearman’s correlation coefficient was used for possible correlations.

The analysis was carried out using SPSS v 29.01 and the significance was set at 0.05 in all cases. Sample matching was done with MatchIt package v. 4.5 using R v4.3 as described in “matching procedure” section, above.

## 3. Results

### Participants

Data for CSF was available for 12 A53T-PD. Consequently, 36 iPD and 36 HC were selected for comparison.

Data for CSF ASSAA was available for 7 A53T-AC; 3/7 had CSF ASSAA assessment at 2 time points.

### Demographic and Clinical features in A53T-PD, i PD and HC

The demographic and other clinical features are summarized in **Table 1**. Age at visit, age at diagnosis of Parkinson’s disease (PD), sex and years of education, MOCA and RBDQS were comparable in all three groups. UPDRS III (OFF), HCY (OFF), GDS and SCOPA -AUT were significantly worse in A53T-PD and i PD compared to HC. UPSIT scores in A53T-PD were significantly lower compared to both i PD and HC.

**Table 1.**
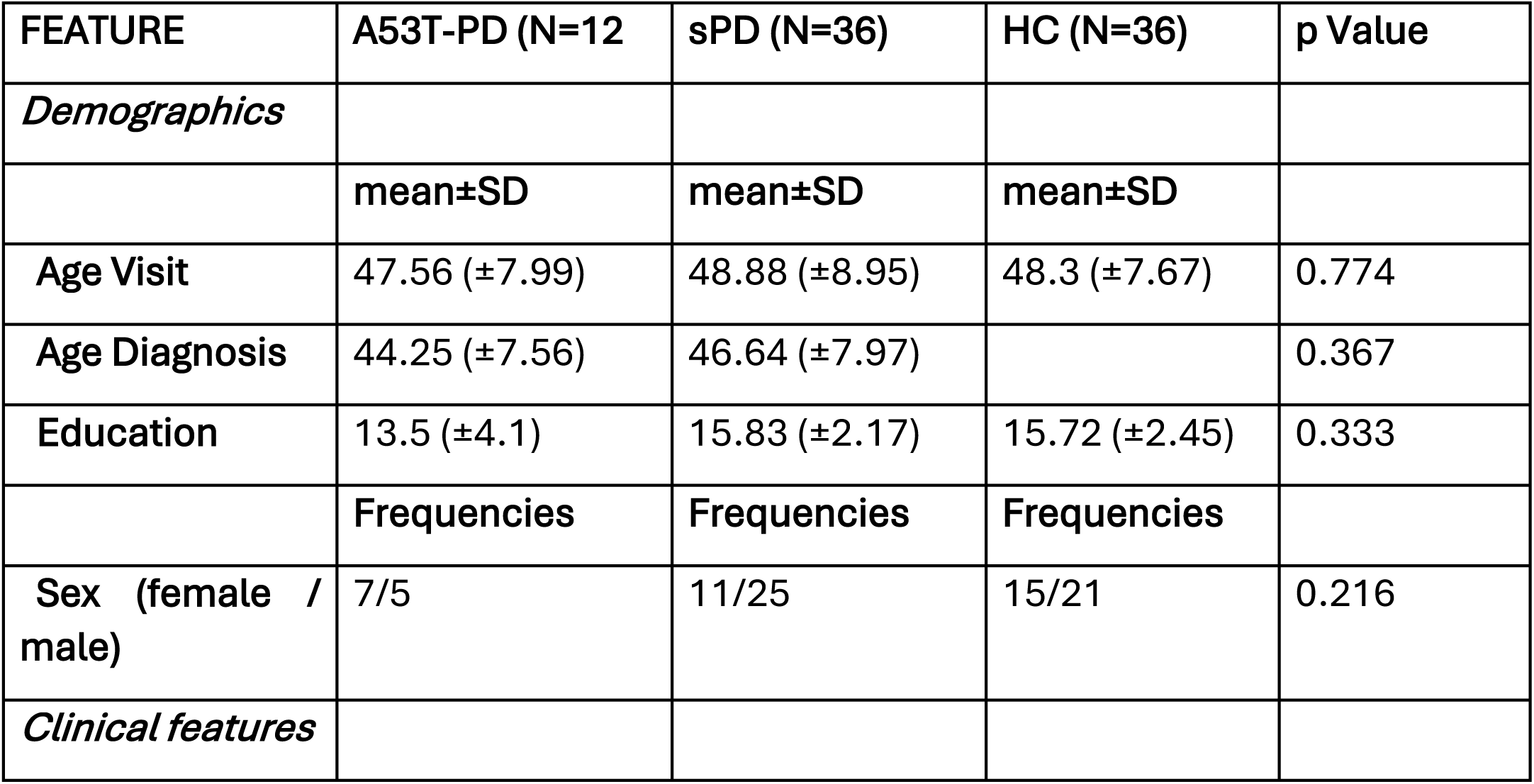
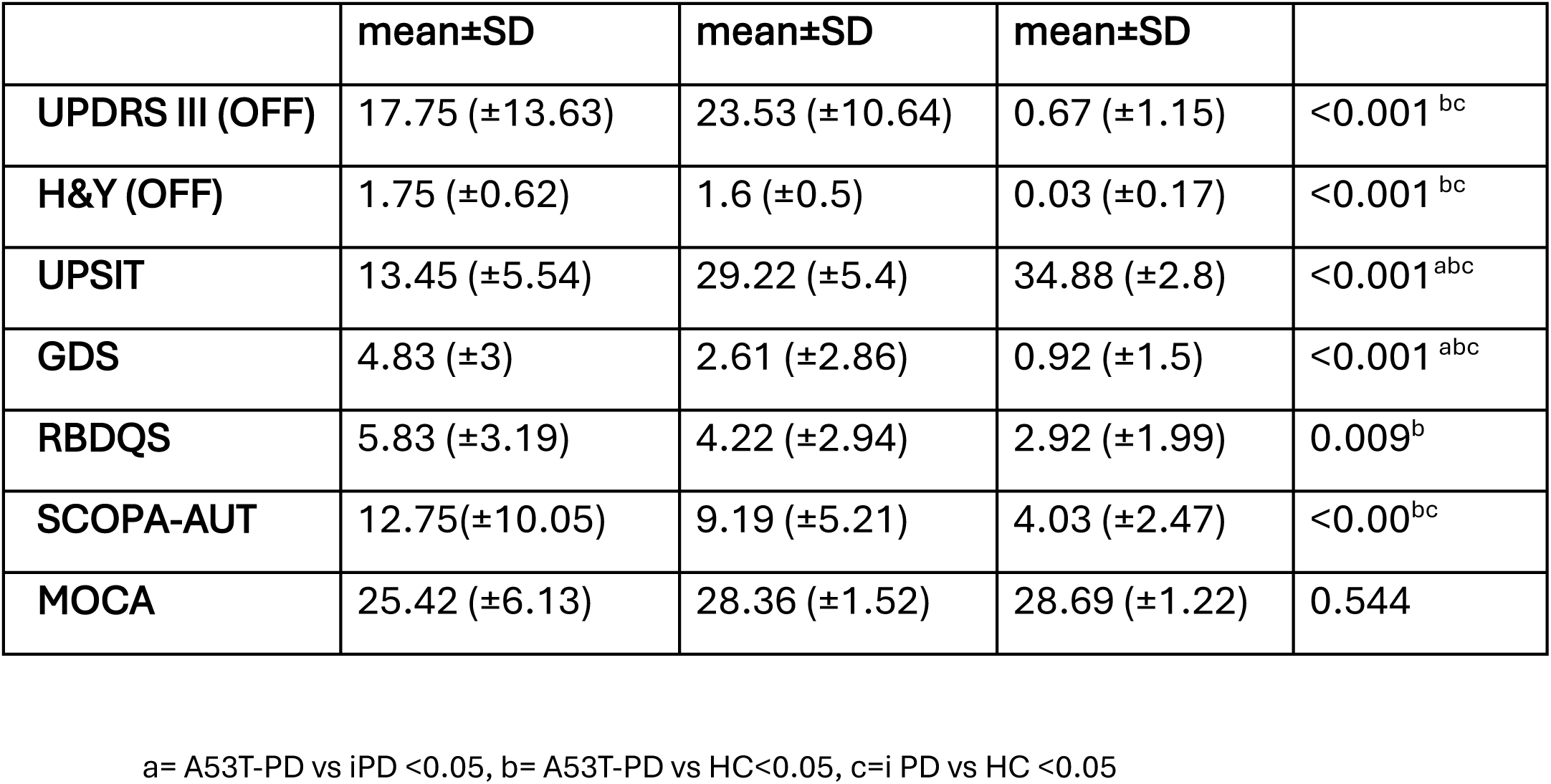
Demographics and Clinical features in A53T-PD, i PD and HC.

### CFS profile in A53T-PD, i PD and HC

All 12 A53T-PD and 33/36 iPD had a positive ASSAA. The other CSF markers were comparable across all groups, although there appeared to be a non-significant trend for lower Abeta 42, t-asyn and p-Tau in the A53T cohort (data shown in **Table 2** and **Figure 1**.). No correlations were found between motor or non-motor features and CSF markers except from a weak positive, statistically significant correlation between t-tau and GDS, t-tau and SCOPA-AUT and between p-tau and GDS in iPD. (data not shown).

**Figure 1:**
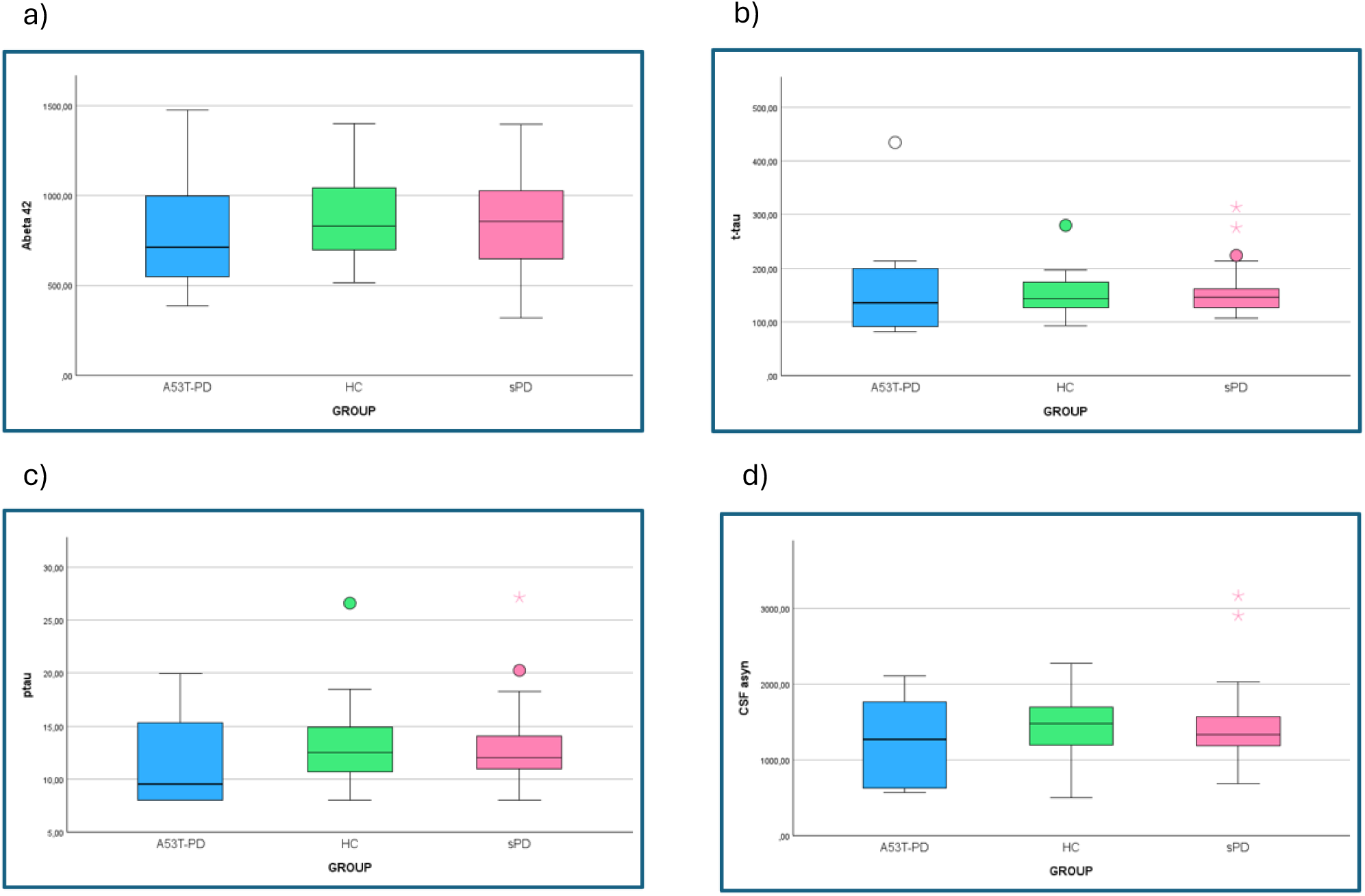

**Table 2.**
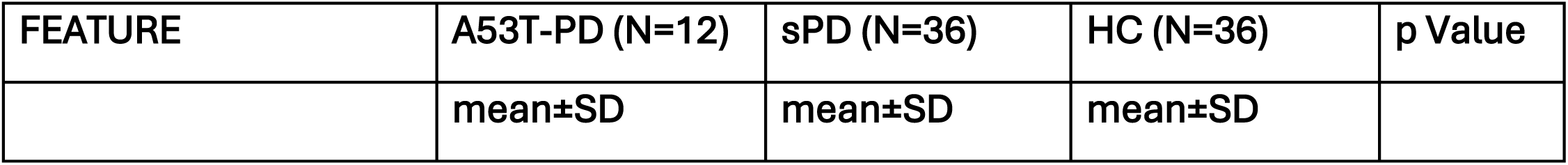
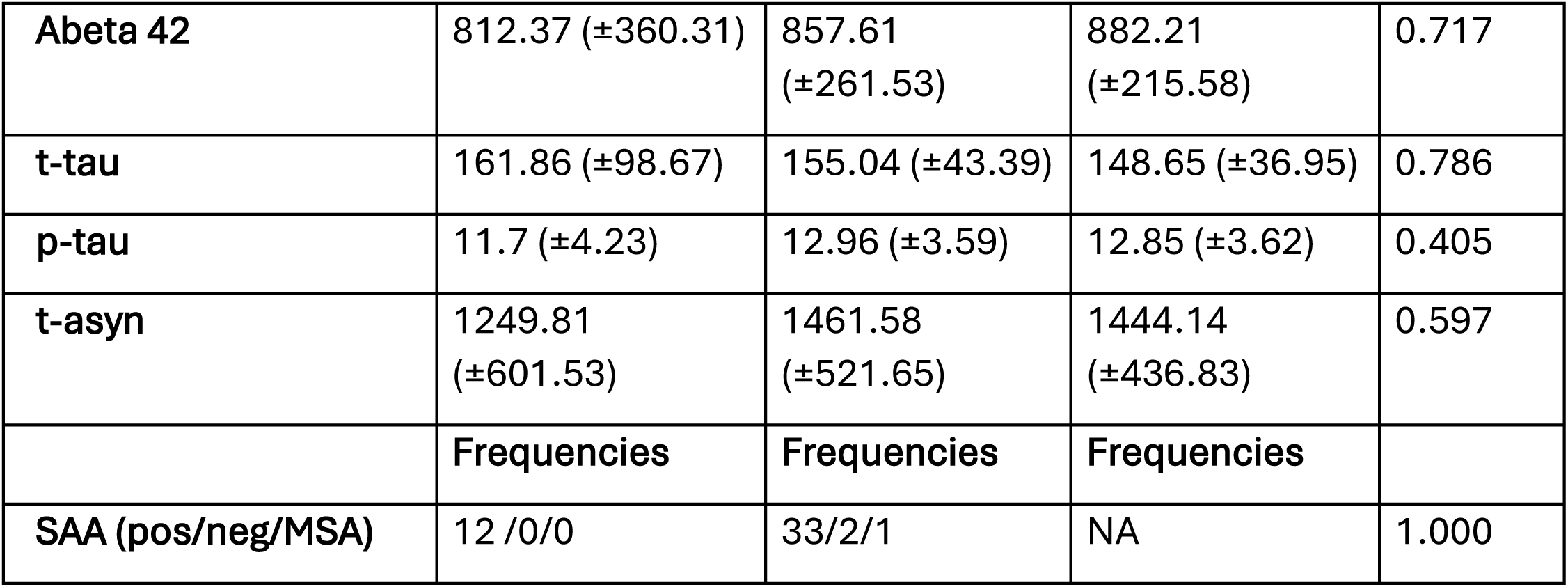
CSF profile in A53T-PD, i PD and HC.

| FEATURE | A53T-PD (N=12) | sPD (N=36) | HC (N=36) | p Value |
| --- | --- | --- | --- | --- |
|  | mean±SD | mean±SD | mean±SD |  |
| **Abeta 42** | 812.37 (±360.31) | 857.61<br>(±261.53) | 882.21<br>(±215.58) | 0.717 |
| **t-tau** | 161.86 (±98.67) | 155.04 (±43.39) | 148.65 (±36.95) | 0.786 |
| **p-tau** | 11.7 (±4.23) | 12.96 (±3.59) | 12.85 (±3.62) | 0.405 |
| **t-asyn** | 1249.81<br>(±601.53) | 1461.58<br>(±521.65) | 1444.14<br>(±436.83) | 0.597 |
|  | **Frequencies** | **Frequencies** | **Frequencies** |  |
| **SAA (pos/neg/MSA)** | 12 /0/0 | 33/2/1 | NA | 1.000 |

### Asymptomatic A53T (A53T-AC)

Data for CSF AS SAA was available for 7 A53T-AC (Table 3):

**Table. 3.** Asymptomatic A53T.

| ID | VISIT<br>PPMI | Y | AGE<br>(range) | SEX | UPSIT | SNIFFIN | MOCA | RBDQ | GDS | SCOPA<br>AUT | H&Y | UPDRS<br>III | AS<br>SAA | abeta | t-tau | p-tau | t-asyn |
| --- | --- | --- | --- | --- | --- | --- | --- | --- | --- | --- | --- | --- | --- | --- | --- | --- | --- |
| 1 | BL | 0 | 61-65 | F | 17 |  | 26 | 1 | 5 | 28 | 2 | 23 | pos |  |  |  |  |
| 2 | BL | 0 | 41-45 | F | 28 |  | 28 | 2 | 0 | 7 | 0 | 0 | neg | 1009 | 165,4 | 13,55 | 1586.7 |
| 2 | V04 | 1 | 46-50 | F |  |  | 29 | 2 | 0 | 2 | 0 | 0 |  |  |  |  |  |
| 2 | V06 | 2 | 46-50 | F |  |  | 29 | 2 | 0 | 2 | 0 | 0 | neg | 1167,6 | 137,4 | 11,42 |  |
| 2 | V08 | 3 | 46-50 | F |  |  | 30 | 3 | 5 | 2 | 0 | 0 |  |  |  |  |  |
| 2 | V10 | 4 | 46-50 | F |  |  | 29 | 2 | 1 | 2 | 0 | 0 |  |  |  |  |  |
| 2 | V12 | 5 | 46-50 | F |  |  | 27 | 1 | 1 | 2 | 0 | 0 |  |  |  |  |  |
| 2 | V14 | 7 | 51-55 | F |  |  | 29 | 1 | 0 | 5 | 0 | 0 |  |  |  |  |  |
| 2 | V15 | 8 | 51-55 | F |  |  | 28 | 1 | 3 | 5 | 0 | 0 |  |  |  |  |  |
| 2 | V18 | 11 | 51-55 | F | 32 |  | 28 | 2 | 1 | 4 | 0 | 0 |  |  |  |  |  |
| 3 | BL | 0 | 41-45 | F | 29 |  | 24 | 5 | 6 | 7 | 0 | 0 | inc | 1475,4 | 214,3 | 17,77 | 2115,5 |
| 3 | V04 | 1 | 41-45 | F |  |  | 27 | 5 | 3 | 5 | 0 | 0 | pos | 1475,4 | 239,1 | 20,29 | 2122,6 |
| 3 | - | 9 | 51-55 | F |  | 5 | 27 | 7 |  | 6 | 0 | 0 |  |  |  |  |  |
| 4 | BL | 0 | 41-45 | F | 32 |  | 28 | 0 | 0 | 1 | 0 | 0 | pos | 1061 | 218,9 | 16,77 | 1802 |
| 4 | V04 | 1 | 41-45 | F |  |  | 28 | 0 | 0 | 2 | 0 | 0 | pos | 1197,2 | 203,5 | 15,63 |  |
| 4 | V06 | 2 | 41-45 | F |  |  | 28 | 1 | 0 | 2 | 0 | 0 |  |  |  |  |  |
| 4 | V08 | 3 | 41-45 | F | 30 |  | 29 | 2 | 0 | 4 | 0 | 0 |  |  |  |  |  |
| 4 | V10 | 4 | 46-50 | F |  |  | 30 | 1 | 0 | 2 | 0 | 0 |  |  |  |  |  |
| 4 | V13 | 6 | 46-50 | F |  |  | 30 | 0 | 0 | 3 | 0 | 0 |  |  |  |  |  |
| 4 | V14 | 7 | 46-50 | F |  |  | 30 | 1 | 0 | 4 | 0 | 0 |  |  |  |  |  |
| 4 | V15 | 8 | 46-50 | F |  |  | 30 | 0 | 0 | 3 | 1 | 8 |  |  |  |  |  |
| 4 | V16 | 9 | 51-55 | F |  |  | 28 | 2 | 0 | 7 | 1 | 16 |  |  |  |  |  |
| 4 | V17 | 10 | 51-55 | F |  |  | 30 | 1 | 0 | 6 | 2 | 6 (ON) |  |  |  |  |  |
| 5 | BL | 0 | 26-30 | M | 34 |  | 30 | 3 | 1 | 1 | 0 | 0 | neg |  |  |  |  |
| 6 | BL | 0 | 26-30 | F |  |  | 30 | 0 | 0 | 0 | 0 | 0 |  |  |  |  |  |
| 6 | V13 | 6 | 31-35 | F |  |  | 29 | 0 | 1 | 1 | 0 | 0 | neg |  |  |  |  |
| 6 | V14 | 7 | 31-35 | F | 36 |  | 28 | 0 | 0 | 0 | 0 | 0 |  |  |  |  |  |
| 7 | BL | 0 | 26-30 | F | 35 |  | 28 | 0 | 3 | 7 | 0 | 0 |  |  |  |  |  |
| 7 | V13 | 6 | 31-35 | F |  |  | 30 | 0 | 2 | 2 | 0 | 0 | pos |  |  |  |  |
| 7 | V14 | 7 | 31-35 | F | 29 |  | 28 | 0 | 1 | 1 | 0 | 0 |  |  |  |  |  |

Y: year, neg: negative, pos: positive, inc: inconclusive

Subject no.1 at BL had a positive CSF AS SAA with motor signs indicating incipient PD; she also presented anosmia (UPSIT: 17), indications of depression (GDS: 5) and a high score in the SCOPA-AUT scale (28) indicating significant autonomic dysfunction.

Subjects 2, 3 and 4 had a CSF AS SAA at two time points:

Subject no.2 had a negative CSF AS SAA and no motor symptoms at two initial time points. At 11 years from BL, the subject did not present any motor or non-motor symptoms or signs except for mild microsmia (UPSIT: 32), which was even improved from BL (UPSIT: 28).

Subject no.3 converted from an inconclusive to a positive CSF AS SAA from the first to the second time point, a year later. This subject was examined 9 years after the positive CSF AS SAA outside the scope of the PPMI study; she remained asymptomatic but had evidence of significant hyposmia (5/12 on Sniffin test). At the baseline visit her UPSIT score was 29, indicating moderate microsmia. She also had some indications of dream enactment behavior and mild autonomic dysfunction already from the baseline visit, but these did not progress over time.

Subject no.4 had a positive CSF AS SAA at BL, confirmed 2 years later; she presented an uneventful course for many years, without motor or non-motor symptoms or signs except for mild microsmia (UPSIT 32 at BL, 30 3 years later). Eight years after BL, she developed unilateral PD motor signs that were progressive, and, a year later, a mild increase of SCOPA-AUT score, suggesting autonomic dysfunction, while cognition remained intact.

Subject no.5 had a negative CSF AS SAA at BL, with no indications of motor or non-motor symptoms or signs. There is no follow-up for this subject.

Subject no.6 had no motor or non-motor symptoms or signs at BL and during a 7-year follow-up, with an intact olfactory function (UPSIT 36) at the last assessment. She had a negative CSF AS SAA 6 years after BL.

Subject no.7 was asymptomatic at BL, with an intact olfactory function (UPSIT: 35). Six years after BL, she tested positive on the CSF AS SAA. One year later, she had moderate microsmia (UPSIT 29), representing a significant drop from BL testing, but no other motor or non-motor symptoms or signs.

## 4. Discussion

Our results revealed that A53T-PD have comparable AD-related neurodegenerative CSF markers to iPD and HC, including Abeta amyloid 1-42, total-Tau and p-Tau. There was a trend for lower Abeta amyloid 1-42 in A53T-PD compared to both other groups, but it was not statistically significant. Notably, these results are based on baseline assessments in a background of relatively intact global cognition in A53T-PD (mean MOCA score of 26, not significantly different from iPD and HC). As we have previously reported, A53T-PD subjects within the first 3-5 years of disease onset maintain intact global cognition, although specific cognitive deficits reflecting frontoparietal network dysfunction emerge ^1, 2^. It is generally only later that global cognitive dysfunction and dementia occur ^2, 12^. In our previous report in a smaller group of A53T-PD subjects, but with more variable disease duration and without a comparator group, we found that 2 subjects with significant global cognitive deficits manifested marginally low Abeta amyloid 1-42 levels^10^. It is therefore possible that A53T-PD subjects may start manifesting alterations in such neurodegenerative markers upon further disease evolution, associated with global cognitive decline, as occurs in iPD^13, 14^, but this hypothesis will require assessment in longitudinal studies. Current data on longitudinal follow-up of such markers within PPMI are scant. Another aspect of note regards levels of total Tau. In our previous report^10^, we had identified two subjects with the peculiar initial manifestation of aggressive Frontotemporal Dementia, followed by rapid cognitive decline and severe brain atrophy, that is uncharacteristic of this genetic cohort. Both these subjects had high levels of total Tau, reflective of the presumed enhanced neurodegeneration. One of these subjects was included in the current PPMI cohort and high total Tau levels (434, 5pg/ml) were confirmed, however this was an exception, and such levels were not high in any other subject or in the overall group of A53T-PD compared to the other two.

Levels of total a-syn were statistically comparable between groups, although there was a definite trend for lower levels in A53T-PD. Levels in iPD were very close to those of HC, whereas in larger cohorts, including the total PPMI cohort^15^, a mild decrease has been noted, presumably reflecting the transition of a-syn to less soluble species due to aggregation. The relatively lower total asyn levels in A53T-PD may reflect an enhanced asyn aggregation process in such subjects, as identified by neuropathology^8^, although the small number assessed precludes definite conclusions.

All A53T-PD had positive ASSAA in CSF, comparable to i PD but unlike other genetic PD. Siderowf et al 2023^16^, found that the proportion of participants with positive α-synuclein SAA results was highest for GBA Parkinson’s disease (95, 9%), followed by sporadic Parkinson’s disease (93, 3%), and lowest for LRRK2 Parkinson’s disease (67·5%). In the same study all participants with positive α-synuclein SAA results that underwent post-mortem examination had typical Lewy pathology, whereas a LRRK2 carrier with preserved olfaction and negative ASSAA had no Lewy pathology. These findings support the correlation of ASSAA positivity and LB pathology and justify the universal positivity of ASSAA in A53T-PD due to severe and widespread LB pathology in this rare genetic synucleinopathy^8^, and despite a report based on A53T SNCA transgenic mice that suggested poor seeding of the WT substrate with this mutant form^11^. This may indicate that a significant proportion of seeds present in the mutant carriers may have a WT conformation, based on the heterozygote nature of the disease.

The results from the CSF AS SAAs in A53T-AC, combined with longitudinal clinical follow-up in most cases, enable a partial recreation of the clinic-pathological evolution of the disease at this pre-symptomatic stage. One A53T-AC (subject no.1) had at baseline a positive CSF AS SAA with motor signs indicating PD and a plethora of non-motor symptoms and signs, therefore should rather be classified as incipient PD and not asymptomatic, but this was the designation in the PPMI data base. For this reason, this subject will no longer be considered in the discussion of asymptomatic carriers. Subjects 2, 5 and 6 are not very informative, as they had negative CSF AS SAAs and uneventful clinical courses. Nevertheless, these results do indicate that the mutation status can be associated with a negative CSF AS SAA in the late 40s, as in subject 2, beyond the mean age of onset of the disease^12^. It will be interesting to have repeat CSF studies at later ages in such individuals, who may even escape the disease, as the p.A53T SNCA mutation is not fully penetrant^12^.

The more relevant discussion centers on subjects 3, 4 and 7. Subject 4, while completely asymptomatic, had a positive CSF AS SAA at BL, confirmed a year later. She manifested stable and acceptable olfactory function at initial visits, and no other non-motor manifestations, indicating that the CSF AS SAA became positive even before olfactory decline. This subject eventually, in a 8-year follow-up after a positive CSF AS SAA, phenoconverted to unilateral motor PD, which progressed, while slight autonomic dysfunction emerged a year later. No UPSIT testing was performed after initial visits, and therefore the evolution of possible olfactory dysfunction could not be ascertained. RBDSQ scores remained low throughout the disease course. Subject 3, while remaining asymptomatic for 9 years after CSF AS SAA conversion from inconclusive to positive, manifested markedly reduced olfactory perception at this time point. Smoldering modest scores in RBDSQ and SCOPA-AUT over this period are unlikely to represent disease manifestations, as they did not progress over time. This subject, at around the time of CSF AS SAA conversion, had acceptable olfactory function and, notably, a polysomnography study that was negative for RBD^3^, reflecting the often quoted lack of specificity of such questionnaires for RBD diagnosis in PD. This is also consistent with the finding that RBD is not generally detected by polysomnography in A53T-AC^3^, although it is reported retrospectively from most A53T-PD subjects, with a temporal onset close to motor symptoms^12^. Therefore, this subject at the time of CSF AS SAA positivity remained without RBD or significant olfactory dysfunction, while hyposmia emerged only later. Subject no.7 has manifested no motor or non-motor symptoms or signs except for moderate microsmia of unknown initiation relative to CSF AS SAA positivity. Again, in this case CSF AS SAA positivity manifested in the absence of significant olfactory dysfunction, although there was a decline from baseline 6 years earlier. Cognition was not affected in A53T-AC at any time point.

These results overall indicate that, in A53T-AC, CSF AS SAA positivity may be identified before prodromal disease manifestations and many years before motor disease onset, and thus may represent a time point when future disease-modifying interventions may be applied, given the high, albeit incomplete penetrance of the p.A53T SNCA mutation^12^. The identification of a positive CSF AS SAA in the absence of PD prodromal signs is quite unique as an observation and suggests that this biomarker may become positive even before prodromal signs emerge. A small percentage of control cases or asymptomatic carriers of other genetic forms of PD are consistently reported with positive CSF AS SAA, including in the PPMI study^16^, but have not been rigorously assessed for non-motor manifestations at the time of positive testing or for possible future evolution to PD. Systematic longitudinal studies in A53T-AC with careful monitoring of symptomatology and semiology, especially of non-motor symptoms, will be required to shed further light on the temporal course of events leading to even the slightest evidence of phenoconversion after the establishment of CSF AS SAA positivity. In parallel, longitudinal imaging assessments, such as DAT and MIBG, and laboratory-based assessments of sleep and autonomic function could also be performed periodically to provide objective measures of disease manifestations. This will be crucial to the understanding of the very early stages of pathobiology of PD.

## Data Availability

Data used in the preparation of this article were obtained on 2026-05-11 from the Parkinson's Precision Medicine Initiative (PPMI) database (https://www.ppmi-info.org/access-data-specimens/download-data), RRID:SCR_006431. For up-to-date information on the study, visit http://www.ppmi-info.org. This analysis was conducted by the PPMI Statistics Core and used actual dates of activity for participants, a restricted data element not available to public users of PPMI data.

## Conflict of Interests

AMS has received funding from the Michael J Fox Foundation for his participation in the PPMI2 study and the “Longitudinal evaluation of a cohort of asymptomatic SNCA mutation carriers to investigate early events in PD pathobiology” study.

NP is employed by the National and Kapodistrian University of Athens and is a scientific consultant for the European Academy of Neurology and has been funded by Michael J. Fox foundation.

IA has received funding from the Michael J Fox Foundation for her participation in the “Longitudinal evaluation of a cohort of asymptomatic SNCA mutation carriers to investigate early events in PD pathobiology” study.

MP has received speaker honoraria from Bial and Zambon, research grants from the Italian Ministry of Health and the Italian Ministry of University.

PB received consultancies as a member of the advisory board for Zambon, Lundbeck, UCB, Chiesi, Abbvie and Acorda.

DEL reports research support and grants from the Michael J. Fox Foundation (for the Parkinson’s Precision Medicine Initiative) and the National Institutes of Health (NIH) (for the Fibromyalgia Transcutaneous Electrical Nerve Stimulation in Physical Therapy Study [FM-TIPS].

KM received support to his institution (Institute for Neurodegenerative Disorders) from The Michael J Fox Foundation. Consultant for the Michael J Fox Foundation, Mitro, Roche, BMS, J and J, GAIN, Calico, Sanofi, Teva, Biohaven, Merck, GEHC, Lilly, ABLi, and Prothena.

AS has been a consultant to the following companies in the past year: Eli Lilly and Co, Cerevance, CND Life Sciences, BioVie Spark/Roche, Prilenia, Arvinas and Novartis. He is an unpaid advisor to Teitur Therapeutics and Neuropacs. He has served on DSMBs for the Huntington Study Group and The Healey ALS Consortium (Massachusetts General Hospital). He has received grant funding from the Michael J. Fox Foundation and NINDS.

TS has served as a consultant for Blue Rock Therapeutics, Centessa, Critical Path for Parkinsons Consortium (CPP), MJFF, Prevail/Lilly Roche/Genentech, Ventus, Sinopia, Takeda, Vanqua Bio, Ventus, Ventyx, and VIMA Tx. Dr. Simuni has equity in Sinopia and has served on the ad board for AskBio, Biohaven, Booster, GAIN, Janssen, Neuron23, Novartis, Parkinson Study Group, Prevail/Lilly, and Roche/Genentech. Has served as a member of the scientific advisory board of Koneksa and UCB and has received research funding from Neuroderm, Prevail, Roche NINDS, MJFF, and Parkinson’s Foundation.

CK has received funding from the Michael J Fox Foundation for his participation in the PPMI2 study.

LS has received the following grants : PPMI2 (supported by the Michael J. Fox Foundation), “BRAIN PRECISION” (funded by the General Secretariat of Research and Innovation), “Longitudinal evaluation of a cohort of asymptomatic SNCA mutation carriers to investigate early events in PD pathobiology” (funded by the Michael J. Fox Foundation), “MJFF PINK1/PRKN Project» ((site PI, funded by the Michael J. Fox Foundation), “Astroglial ApoE as a mediator of β-amyloid pathology in synucleinopathies” (funded by the Hellenic Foundation for Research and Innvoation), “Targeting the Autophagy Lysosome Pathway in Human MSA” (funded by the MSA Trust, Collaborator), and “CMA as a Means to Counteract alpha-Synuclein Pathology in Non-Human Primates” (funded by the Michael J. Fox Foundation, Collaborator). He has served on Advisory Boards for Abbvie, Innovis Pharma, and ITF Hellas and has received honoraria from ITF Hellas, Innovis Pharma and Abbvie. He has participated in clinical trials as site PI, funded by Roche, AB Science, Immunovant and Sanofi.

## Funding

This research was funded in part by the Write Now award to AMS, in addition to the grant “Longitudinal evaluation of a cohort of asymptomatic SNCA mutation carriers to investigate early events in PD pathobiology” awarded by the MJFF to LS. Eginitio Hospital is a Centre of Excellence of the Rare Disease Network ERN-RND.

## Acknowledgments

Data used in the preparation of this article were obtained on 2026-05-11 from the Parkinson’s Precision Medicine Initiative (PPMI) database (https://www.ppmi-info.org/access-data-specimens/download-data), RRID:SCR_006431. For up-to-date information on the study, visit http://www.ppmi-info.org.

This analysis was conducted by the PPMI Statistics Core and used actual dates of activity for participants, a restricted data element not available to public users of PPMI data.

This study was supported by PPMI, in part by the Write Now award to AMS, in addition to the grant “Longitudinal evaluation of a cohort of asymptomatic SNCA mutation carriers to investigate early events in PD pathobiology” awarded by the MJFF to LS. Eginitio Hospital is a Centre of Excellence of the Rare Disease Network ERN-RND.

PPMI – a public-private partnership – is funded by the Michael J. Fox Foundation for Parkinson’s Research and funding partners, including AbbVie, Alamar Biosciences, Aligning Science Across Parkinson’s (ASAP), Arrowhead Pharma, Arvinas, AskBio, BIAL, BioArctic, Biohaven, BlueRock Therapeutics, Bristol Myers Squibb, Calico Labs, Capsida Biotherapeutics, Critical Path Institute, DaCapo Brainscience, Denali, Edmond J. Safra Foundation, Eli Lilly, Gain Therapeutics, GE Healthcare, Genentech, GSK, Insitro, Johnson C Johnson Innovative Medicine, Lundbeck, Merck, Neumora, Neuron23, Novartis, Olink, Regeneron, Roche, Sanofi, Tenvie, UCB, Vanqua Bio, Voyager Therapeutics, The Weston Family Foundation.

Protocol information for The Parkinson’s Precision Medicine Initiative (PPMI) Clinical - Establishing a Deeply Phenotyped PD Cohort can be found on protocols.io or by following this link: https://dx.doi.org/10.17504/protocols.io.n92ldmw6ol5b/v2.

Statistical analysis codes used to perform the analyses in this article are shared on Zenodo [ DOI: 10.5281/zenodo.17187364 ].

## Appendix

PPMI STUDY TEAMS/CORES/COLLABORATORS FOR PUBLICATIONS

### Executive Steering Committee

Marek, MD1 (Principal Investigator); Caroline Tanner, MD, PhD9; Tanya Simuni, MD3; Andrew Siderowf, MD, MSCE12; Douglas Galasko, MD27; Lana Chahine, MD41; Christopher Coffey, PhD4; Kalpana Merchant, PhD61; Kathleen Poston, MD40; Roseanne Dobkin, PhD43; Tatiana Foroud, PhD15; Brit Mollenhauer, MD8; Dan Weintraub, MD12; Ethan Brown, MD9; Karl Kieburtz, MD, MPH23; Mark Frasier, PhD6; Todd Sherer, PhD6; Sohini Chowdhury, MA6; Roy Alcalay, MD36 and Aleksandar Videnovic, MD47

### Steering Committee

Duygu Tosun-Turgut, PhD9; Werner Poewe, MD7; Susan Bressman, MD14; Jan Hammer15; Raymond James, RN22; Ekemini Riley, PhD42; John Seibyl, MD1; Leslie Shaw, PhD12; David Standaert, MD, PhD18; Sneha Mantri, MD, MS62; Nabila Dahodwala, MD12; Michael Schwarzschild47; Connie Marras45; Hubert Fernandez, MD25; Ira Shoulson, MD23; Helen Rowbotham2; Paola Casalin11 and Claudia Trenkwalder, MD8

**Michael J. Fox Foundation (Sponsor):** Todd Sherer, PhD; Sohini Chowdhury, MA; Mark Frasier, PhD; Jamie Eberling, PhD; Katie Kopil, PhD; Alyssa O’Grady; Maggie McGuire Kuhl; Leslie Kirsch, EdD and Tawny Willson, MBS

### Site Investigators

Charles Adler, PhD51; Roy Alcalay, MD36; Amy Amara, PhD52; Paolo Barone, PhD30; Bastiaan Bloem, PhD60 Susan Bressman, MD14; Kathrin Brockmann, MD26; Norbert Brüggemann, MD59; Lana Chahine, MD41; Kelvin Chou, MD44; Nabila Dahodwala, MD12; Alberto Espay, MD32; Stewart Factor, DO16; Hubert Fernandez, MD25; Michelle Fullard, MD52; Douglas Galasko, MD27; Robert Hauser, MD19; Penelope Hogarth, MD17; Shu-Ching Hu, PhD21; Michele Hu, PhD 58; Stuart Isaacson, MD31; Christine Klein, MD59; Rejko Krueger, MD2; Mark Lew, MD49; Zoltan Mari, MD56; Connie Marras, PhD45; Maria Jose Martí, PhD 34; Nikolaus McFarland, PhD54; Tiago Mestre, PhD46; Brit Mollenhauer, MD 8; Emile Moukheiber, MD28; Alastair Noyce, PhD63 Wolfgang Oertel, PhD64; Njideka Okubadejo, MD65; Sarah O’Shea, MD39; Rajesh Pahwa, MD48; Nicola Pavese, PhD57; Werner Poewe, MD7; Ron Postuma, MD55; Giulietta Riboldi, MD53; Lauren Ruffrage, MS18; Javier Ruiz Martinez, PhD 35; David Russell, PhD1; Marie H Saint-Hilaire, MD22; Neil Santos, BS51; Wesley Schlett47; Ruth Schneider, MD23; Holly Shill, MD50; David Shprecher, DO24; Tanya Simuni, MD3; David Standaert, PhD18; Leonidas Stefanis, PhD38; Yen Tai, PhD29; Caroline Tanner, PhD9; Arjun Tarakad, MD20; Eduardo Tolosa PhD34 and Aleksandar Videnovic, MD47

### Coordinators

Susan Ainscough, BA30; Courtney Blair, MA18; Erica Botting19; Isabella Chung, BS56; Kelly Clark24; Ioana Croitoru35; Kelly DeLano, MS32; Iris Egner, PhD7; Fahrial Esha, BS53; May Eshel36; Frank Ferrari, BS44; Victoria Kate Foster57; Alicia Garrido, MD34; Madita Grümmer59; Bethzaida Herrera50; Ella Hilt26; Chloe Huntzinger, BA52; Raymond James, BS22; Farah Kausar, PhD9; Christos Koros, MD, PhD38; Yara Krasowski60; Dustin Le, BS 17; Ying Liu, MD52; Taina M. Marques, PhD2; Helen Mejia Santana, MA39; Sherri Mosovsky, MPH41; Jennifer Mule, BS25; Philip Ng, BS45; Lauren O’Brien48; Abiola Ogunleye, PGDip29; Oluwadamilola Ojo, MD65; Obi Onyinanya, BS28; Lisbeth Pennente, BA31; Romina Perrotti55; Michael Pileggi, MS55; Ashwini Ramachandran, MSc12; Deborah Raymond, MS14; Jamil Razzaque, MS58; Shawna Reddie, BA46; Kori Ribb, BSN, 28; Kyle Rizer, BA54; Janelle Rodriguez, BS27; Stephanie Roman, HS1; Clarissa Sanchez, MPH20; Cristina Simonet, PhD29; Anisha Singh, BS23; Elisabeth Sittig64; Barbara Sommerfeld MSN16; Angela Stovall, BS 44; Bobbie Stubbeman, BS 32; Alejandra

Valenzuela, BS49; Catherine Wandell, BS21; Diana Willeke8; Karen Williams, BA3 and Dilinuer Wubuli, MB45

1. Institute for Neurodegenerative Disorders, New Haven, CT
2. University of Luxembourg, Luxembourg
3. Northwestern University, Chicago, IL
4. University of Iowa, Iowa City, IA
5. VectivBio AG
6. The Michael J. Fox Foundation for Parkinson’s Research, New York, NY
7. Innsbruck Medical University, Innsbruck, Austria
8. Paracelsus-Elena Klinik, Kassel, Germany
9. University of California, San Francisco, CA
10. Laboratory of Neuroimaging (LONI), University of Southern California
11. BioRep, Milan, Italy
12. University of Pennsylvania, Philadelphia, PA
13. National Institute on Aging, NIH, Bethesda, MD
14. Mount Sinai Beth Israel, New York, NY
15. Indiana University, Indianapolis, IN
16. Emory University of Medicine, Atlanta, GA
17. Oregon Health and Science University, Portland, OR
18. University of Alabama at Birmingham, Birmingham, AL
19. University of South Florida, Tampa, FL
20. Baylor College of Medicine, Houston, TX
21. University of Washington, Seattle, WA
22. Boston University, Boston, MA
23. University of Rochester, Rochester, NY
24. Banner Research Institute, Sun City, AZ
25. Cleveland Clinic, Cleveland, OH
26. University of Tübingen, Tübingen, Germany
27. University of California, San Diego, CA
28. Johns Hopkins University, Baltimore, MD
29. Imperial College of London, London, UK
30. University of Salerno, Salerno, Italy
31. Parkinson’s Disease and Movement Disorders Center, Boca Raton, FL
32. University of Cincinnati, Cincinnati, OH
33. Hospital Clinic of Barcelona, Barcelona, Spain
34. Hospital Universitario Donostia, San Sebastian, Spain
35. Tel Aviv Sourasky Medical Center, Tel Aviv, Israel
36. St. Olav’s University Hospital, Trondheim, Norway
37. National and Kapodistrian University of Athens, Athens, Greece
38. Columbia University Irving Medical Center, New York, NY
39. Stanford University, Stanford, CA
40. University of Pittsburgh, Pittsburgh, PA
41. Center for Strategy Philanthropy at Milken Institute, Washington D.C.
42. 12, New Brunswick, NJ
43. University of Michigan, Ann Arbor, MI
44. Toronto Western Hospital, Toronto, Canada
45. The Ottawa Hospital, Ottawa, Canada
46. Massachusetts General Hospital, Boston, MA
47. University of Kansas Medical Center, Kansas City, KS
48. University of Southern California, Los Angeles, CA
49. Barrow Neurological Institute, Phoenix, AZ
50. Mayo Clinic Arizona, Scottsdale, AZ
51. University of Colorado, Aurora, CO
52. NYU Langone Medical Center, New York, NY
53. University of Florida, Gainesville, FL
54. Montreal Neurological Institute and Hospital/McGill, Montreal, QC, Canada
55. Cleveland Clinic-Las Vegas Lou Ruvo Center for Brain Health, Las Vegas, NV
56. Clinical Ageing Research Unit, Newcastle, UK
57. John Radcliffe Hospital Oxford and Oxford University, Oxford, UK
58. Universität Lübeck, Luebeck, Germany
59. Radboud University, Nijmegen, Netherlands
60. TransThera Consulting
61. Duke University, Durham, NC
62. Wolfson Institute of Population Health, Queen Mary University of London, UK
63. Philipps-University Marburg, Germany
64. University of Lagos, Nigeria

## Notes

### Author Declarations

The present study was conducted in agreement with the principles of the Declaration of Helsinki. The Scientific Board of all PPMI sites involved gave ethical approval for this work.

## References

1. Koros C, Stamelou M, Simitsi A, Beratis I, Papadimitriou D, Papagiannakis N, Fragkiadaki S, Kontaxopoulou D, Papageorgiou SG, Stefanis L. Selective cognitive impairment and hyposmia in p.A53T SNCA PD vs typical PD. Neurology. 2018 Mar 6;90(10):e864–e869. doi: 10.1212/WNL.0000000000005063. Epub 2018 Feb 7. PMID: 29438043.

2. Simitsi AM, Sfikas E, Koros C, Papagiannakis N, Beratis I, Papadimitriou D, Antonellou R, Fragiadaki S, Kontaxopoulou D, Picillo M, Pachi I, Alefanti I, Stamelou M, Barone P, Stefanis L. Motor and nonmotor features of p.A53T alpha-synuclein PD vs idiopathic PD: longitudinal data from the PPMI study. J Neurol. 2025 Feb 12;272(3):203. doi: 10.1007/s00415-024-12836-w. PMID: 39934455; PMCID: PMC11814018.

3. Simitsi AM, Koros C, Stamelou M, Papadimitriou D, Leonardos A, Bougea A, Papagiannakis N, Pachi I, Angelopoulou E, Lourentzos K, Bonakis A, Stefanis L. REM sleep behavior disorder and other sleep abnormalities in p. A53T SNCA mutation carriers. Sleep. 2021 May 14;44(5):zsaa248. doi: 10.1093/sleep/zsaa248. PMID: 33231251.

4. Spillantini MG, Schmidt ML, Lee VM, Trojanowski JQ, Jakes R, Goedert M. Alpha-synuclein in Lewy bodies. Nature. 1997 Aug 28;388(6645):839–40. doi: 10.1038/42166. PMID: 9278044.

5. Pramstaller PP, Schlossmacher MG, Jacques TS, Scaravilli F, Eskelson C, Pepivani I, Hedrich K, Adel S, Gonzales-McNeal M, Hilker R, Kramer PL, Klein C. Lewy body Parkinson’s disease in a large pedigree with 77 Parkin mutation carriers. Ann Neurol. 2005 Sep;58(3):411–22. doi: 10.1002/ana.20587. PMID: 16130111.

6. Ross OA, Toft M, Whittle AJ, Johnson JL, Papapetropoulos S, Mash DC, Litvan I, Gordon MF, Wszolek ZK, Farrer MJ, Dickson DW. Lrrk2 and Lewy body disease. Ann Neurol. 2006 Feb;59(2):388–93. doi: 10.1002/ana.20731. PMID: 16437559.

7. Kalia LV, Lang AE, Hazrati LN, Fujioka S, Wszolek ZK, Dickson DW, Ross OA, Van Deerlin VM, Trojanowski JQ, Hurtig HI, Alcalay RN, Marder KS, Clark LN, Gaig C, Tolosa E, Ruiz-Martínez J, Marti-Masso JF, Ferrer I, López de Munain A, Goldman SM, Schüle B, Langston JW, Aasly JO, Giordana MT, Bonifati V, Puschmann A, Canesi M, Pezzoli G, Maues De Paula A, Hasegawa K, Duyckaerts C, Brice A, Stoessl AJ, Marras C. Clinical correlations with Lewy body pathology in LRRK2-related Parkinson disease. JAMA Neurol. 2015 Jan;72(1):100–5. doi: 10.1001/jamaneurol.2014.2704. PMID: 25401511; PMCID: PMC4399368.

8. Spira PJ, Sharpe DM, Halliday G, Cavanagh J, Nicholson GA. Clinical and pathological features of a Parkinsonian syndrome in a family with an Ala53Thr alpha-synuclein mutation. Ann Neurol. 2001 Mar;49(3):313–9. PMID: 11261505.

9. Simuni T, Chahine LM, Poston K, Brumm M, Buracchio T, Campbell M, Chowdhury S, Coffey C, Concha-Marambio L, Dam T, DiBiaso P, Foroud T, Frasier M, Gochanour C, Jennings D, Kieburtz K, Kopil CM, Merchant K, Mollenhauer B, Montine T, Nudelman K, Pagano G, Seibyl J, Sherer T, Singleton A, Stephenson D, Stern M, Soto C, Tanner CM, Tolosa E, Weintraub D, Xiao Y, Siderowf A, Dunn B, Marek K. A biological definition of neuronal α-synuclein disease: towards an integrated staging system for research. Lancet Neurol. 2024 Feb;23(2):178–190. doi: 10.1016/S1474-4422(23)00405-2. PMID: 38267190.

10. Bougea A, Koros C, Stamelou M, Simitsi A, Papagiannakis N, Antonelou R, Papadimitriou D, Breza M, Tasios K, Fragkiadaki S, Geronicola Trapali X, Bourbouli M, Koutsis G, Papageorgiou SG, Kapaki E, Paraskevas GP, Stefanis L. Frontotemporal dementia as the presenting phenotype of p.A53T mutation carriers in the alpha-synuclein gene. Parkinsonism Relat Disord. 2017 Feb;35:82–87. doi: 10.1016/j.parkreldis.2016.12.002. Epub 2016 Dec 6. PMID: 28012952.

11. Han JY, Jang HS, Green AJE, Choi YP. RT-QuIC-based detection of alpha-synuclein seeding activity in brains of dementia with Lewy Body patients and of a transgenic mouse model of synucleinopathy. Prion. 2020 Dec;14(1):88–94. doi: 10.1080/19336896.2020.1724608. PMID: 32041499; PMCID: PMC7039666.

12. Papadimitriou D, Antonelou R, Miligkos M, Maniati M, Papagiannakis N, Bostantjopoulou S, Leonardos A, Koros C, Simitsi A, Papageorgiou SG, Kapaki E, Alcalay RN, Papadimitriou A, Athanassiadou A, Stamelou M, Stefanis L. Motor and Nonmotor Features of Carriers of the p.A53T Alpha-Synuclein Mutation: A Longitudinal Study. Mov Disord. 2016 Aug;31(8):1226–30. doi: 10.1002/mds.26615. Epub 2016 Mar 29. PMID: 27028329.

13. Siderowf A, Xie SX, Hurtig H, Weintraub D, Duda J, Chen-Plotkin A, Shaw LM, Van Deerlin V, Trojanowski JQ, Clark C. CSF amyloid {beta} 1-42 predicts cognitive decline in Parkinson disease. Neurology. 2010 Sep 21;75(12):1055–61. doi: 10.1212/WNL.0b013e3181f39a78. Epub 2010 Aug 18. PMID: 20720189; PMCID: PMC2942062.

14. Schrag A, Siddiqui UF, Anastasiou Z, Weintraub D, Schott JM. Clinical variables and biomarkers in prediction of cognitive impairment in patients with newly diagnosed Parkinson’s disease: a cohort study. Lancet Neurol. 2017 Jan;16(1):66–75. doi: 10.1016/S1474-4422(16)30328-3. Epub 2016 Nov 18. PMID: 27866858; PMCID: PMC5377592.

15. Kang JH, Mollenhauer B, Coffey CS, Toledo JB, Weintraub D, Galasko DR, Irwin DJ, Van Deerlin V, Chen-Plotkin AS, Caspell-Garcia C, Waligórska T, Taylor P, Shah N, Pan S, Zero P, Frasier M, Marek K, Kieburtz K, Jennings D, Tanner CM, Simuni T, Singleton A, Toga AW, Chowdhury S, Trojanowski JQ, Shaw LM; Parkinson’s Progression Marker Initiative. CSF biomarkers associated with disease heterogeneity in early Parkinson’s disease: the Parkinson’s Progression Markers Initiative study. Acta Neuropathol. 2016 Jun;131(6):935–49. doi: 10.1007/s00401-016-1552-2. Epub 2016 Mar 28. PMID: 27021906; PMCID: PMC5031365

16. Siderowf A, Concha-Marambio L, Lafontant DE, Farris CM, Ma Y, Urenia PA, Nguyen H, Alcalay RN, Chahine LM, Foroud T, Galasko D, Kieburtz K, Merchant K, Mollenhauer B, Poston KL, Seibyl J, Simuni T, Tanner CM, Weintraub D, Videnovic A, Choi SH, Kurth R, Caspell-Garcia C, Coffey CS, Frasier M, Oliveira LMA, Hutten SJ, Sherer T, Marek K, Soto C; Parkinson’s Progression Markers Initiative. Assessment of heterogeneity among participants in the Parkinson’s Progression Markers Initiative cohort using α-synuclein seed amplification: a cross-sectional study. Lancet Neurol. 2023 May;22(5):407–417. doi: 10.1016/S1474-4422(23)00109-6. PMID: 37059509; PMCID: PMC10627170.

